# An Adjunct Role Of Potassium Rich Vegetarian Diet And A Novel Potassium Food Supplement To Improve Pain In Chronic Rheumatoid Arthritis On Supervised Standard Care: A Randomized Controlled Study

**DOI:** 10.1101/2022.06.24.22276843

**Authors:** Kianifard Toktam, Saluja Manjit, Sarmukaddam Sanjeev, Venugopalan Anuradha, Chopra Arvind

**Affiliations:** Primary Care Dietitians, UK; Center for Rheumatic Diseases, Pune, India

**Keywords:** Potassium, Diet, Rheumatoid arthritis, Pain, Vegetarian Diet

## Abstract

**Introduction:** Potassium inadequacy (diet and body storage) may adversely affect rheumatoid arthritis (RA) and is sparsely reported. We evaluated the therapeutic benefits (RA) of food-based potassium intake, recommended daily allowance (RDA), and higher.

**Objective:** To evaluate pain reduction by oral potassium in chronic RA

**Methods:** 172 consenting and eligible symptomatic patients (median duration 6.5 years) on ongoing standard care were randomized in a single-center study (80% power, significant p < 0.05) - Arm A (vegetarian diet as per the India RDA for potassium), Arm B (Arm A diet plus novel potassium food supplement) and Arm C (regular diet, control). Efficacy and safety, and diet intake (three-day recall, Food Composition tables) were assessed (blinded) at monthly intervals till 16-week of study completion and statistically analyzed using standard methods. Study groups were found matched and showed inadequate baseline dietary potassium (RDA). On study completion, the median daily potassium intake was 2959 mg in Arm A, 6063 mg in Arm B, and 2553 mg in Arm C. Study subjects remained normokalemic at all evaluations. Overall, the background medication remained stable.

**Results:** 155 patients (90.1%) completed the study. Adverse events were mild. On comparison, the improvement in pain (primary efficacy) on study completion was significant in Arm B as per protocol analysis; the mean change in pain visual analog scale from baseline was −2.23 (95% confidence interval −2.99 to −1.48). Arm B showed impressive improvement in joint function. High potassium intake predicted low pain (Likelihood ratio 2.9, logistic regression). Compliance (intervention), diet recall, medication, complex nature of dietary intervention/other nutrients, and lack of placebo were potential confounders to ascertain the effectiveness of potassium.

**Conclusion:** A planned vegetarian diet and food supplement intervention with a predominantly increased potassium intake significantly reduced chronic RA pain. This adjunct treatment was found safe and well tolerated. However, it requires further validation.

**Trial Registration:** Clinical Trial Registry of India- CTRI/2022/03/040726

**KEY MESSAGE:** *What is already known on the topic?:*

1. RA is predominantly managed with drug therapy and diet is often neglected
2. RA is complicated by hypertension and other cardiovascular disorders, and osteoporosis which may benefit from potassium intervention.
3. Potassium and potassium ion channels are important the pathophysiology of pain (and probably inflammation)
4. Patients of RA may be deficient in potassium due to inadequate diet or sarcopenia

*What does this study add?:*

1. Potassium-rich vegetarian diet and a novel high-potassium food supplement significantly reduced pain in chronic RA on supervised standard drug care.
2. Several participants showed improved joint function and better blood pressure status
3. Higher potassium intake based on food and diet was safe and well tolerated

*How this study might affect research, practice, or policy?:*

1. Potassium rich predominantly vegetarian diet should be advocated in the management of RA as an adjuvant
2. A judicious use of high potassium food supplement along with suitable diet may benefit difficult and chronic RA
3. The current guidelines on oral potassium intake in RA and other medical disorders need to be revised and call for more research

## INTRODUCTION

Rheumatoid arthritis (RA) is a lifelong autoimmune disease characterized by painful polyarthritis and articular deformities, and several systemic complications,[1]. RA causes a reduction in productivity, quality of life, and longevity,[1]. There is no cure and the overarching therapeutic target is a remission or low disease activity,[2]. Standard treatment guidelines of the American College of Rheumatology (ACR) are loaded with drugs that are potentially toxic and need close monitoring, and adjuvant therapies like diet are neglected,[3]. Diet has been pivotal to treat RA since ancient times,[4]. But modern evidence in favor of diet in RA seems limited,[5]. The global burden of RA is enormous and often not matched by rheumatology care and services,[6].

Potassium is an essential micronutrient but its role in RA is not yet elucidated,[7–8]. There are sparse reports of potassium deficiency in RA,[7]. However, the consequence of the latter is not known. Potassium deficiency may also cause general body aches and pains and debility which are common features in RA,[7, 9]. Several decades ago, Weber advocated an etiological link (RA),[10]. Recently, we reported significantly low potassium intake in RA,[11]. High potassium chloride (mixed in grape juice) reduced pain and arthritis in a randomized controlled exploratory study,[12].

The primary objective of the current study was to investigate the adjuvant therapeutic role of potassium to reduce pain in chronic RA. We used a pre-planned potassium-rich vegetarian diet (PRVD) and a novel potassium food supplement (PFS).

## METHODS AND PATIENTS

This non-commercial investigator-initiated study was part of a Ph.D. dissertation,[13]. It was approved by the local (CRD) ethics committee. The protocol adhered to the tenets of the ‘Helsinki Declaration’ (updated) and GCP/ICH, and national guidelines,[14]. The protocol was registered with the Drug Trial Registry of India (CTRI/2022/03/040726),[15]. The study was begun in Jan 2014 and completed in Aug 2015. The period of recruitment was from 12 Feb 2014 to 02 July 2014. The last patient last visit was on 12 Dec 2014.

No changes were made to methods and protocol after commencement of the trial.

### Study Design

This was a randomized, single-blind (assessor), active control, parallel efficacy, drug trial study of 16 weeks duration. It was carried out in a single center (CRD). There were 3 treatment arms-Arm A (PRVD), Arm B (PRVD plus PFS), and Arm C (control routine diet). Following enrollment, each participant was examined at a randomized baseline and thereafter at monthly intervals as per the protocol.

### Participants and Selection

Interested patients from CRD outpatient signed informed consent, and were screened in a fasting state and reviewed within the next 48 hours and randomized if found eligible [15]. Eligible patients suffered from previous painful arthritis of at least 8 weeks duration and received supervised standard care for at least 16 weeks. Study participants were investigated free of cost and received a modest monetary allowance.

Inclusion criteria included (i) adult patients of RA[16] (ii) disease duration of at least 6 months (iii) maximum pain (joints) ≥ 4 cms on a visual analog scale (VAS) in the previous 24 hours (screening). Exclusion criteria included (i) a daily dose of 10 mg or more prednisolone anytime during the previous four weeks (ii) abnormal serum creatinine, blood urea nitrogen assay, and serum potassium (≥ 5.5 mEq/L) assay (iii) any medication known to affect body potassium.

### Efficacy

The primary efficacy was an improvement in the pain on study completion. There were several secondary efficacy measures of disease activity and function.

### Procedures

#### Pre-Randomization Counselling

Randomized participants were counseled and received a study brochure in the local language.

#### Randomization/Enrolment

A standard computer-generated randomization schedule (1:1:1) was prepared by MS (coordinator) and used on a first come first serve basis. Participants were immediately allocated their study arm and study begun under supervision of MS.

#### Blinding

Independent rheumatologists assessed efficacy and adverse events (AE) in a blinded manner.

#### Diet food record

A retrospective three-day diet record, immediately prior to the study visit, was completed in a face-to-face interview by the dietitian (TK) using a validated questionnaire and standard household measures,[11].

#### Clinical

Participants satisfied the 1987 ACR classification criteria (RA),[1,3]. Efficacy measures included a 68/66 joint count for pain/tenderness and swelling (ACR recommendation), a pain visual analog scale (a 10 cm horizontal line anchored at 0 for no pain and 10 for optimum pain) to measure pain, patient and physician global assessment (categorical scale: 1 for asymptomatic and 5 for very severe), RA pain scale, Indian modified Stanford health assessment questionnaire/HAQ to assess joint function (score range 0-24 with a higher score indicating more difficulty), Short form/SF 36-mental and physical (with permission from the vendor) to assess the quality of life (higher score meant better health),[17–20]. Improvement and RA disease activity were assessed by standard indices-Disease Activity Score 28 (DAS 28) and ACR improvement (percent) index (ACR 20 or ACR 70), [21]. Efficacy measures were assessed at monthly interval till study completion.

#### Other

Standard laboratory and radiological investigations for the management of RA were carried out with emphasis on monitoring medication and safety. Serum cortisol and spot urinary potassium and sodium assay were carried out.

### Study Intervention

PRVD and PFS were active interventions (Arm A and Arm B) while a routine diet was the control (Arm C). There was no placebo. Participants were allowed to follow the traditional cooking style and routine meal timings. Participants were not to fast and no other diet was allowed.

#### Diet/Supplement Target

The PRVD provided at least 3500 mg of elemental potassium daily according to Indian guidelines for recommended daily allowance (RDA, 3225 mg for women, and 3750 mg for men),[22–23]. The addition of the PFS in Arm B increased the daily potassium intake to about 5.5 gm daily.

#### Special Diet Brochure

A brochure guided the selection of PRVD food items for daily meals. These items were listed in 4 categories (vegetables, fruits, pulses, and cereals) with multiple choices. Household measures were used to describe individual consumption daily; proportionately increased for a family meal. Excess frying and oil were discouraged. The brochure described the consumption of salt and spices, water, and milk (and dairy products). Non-vegetarian food was discouraged. The brochure was based on local customs and traditions, and affordability.

#### Potassium Food Supplement/PFS (Arm B)

One hundred and forty-two grams (1 unit) of supplement contained green gram (V radiate, 25 gm), cowpea (V unguiculata, 25 gm, coriander seed (C sativum, 25 gm), cumin seed (C cyminum, 25 gm) and 42 gm of oral rehydration salt (Indian pharmacopeia, 3.5 gm sodium chloride, 1.5 gm potassium chloride 2.9 gm trisodium citrate, 13.5 gm glucose (dextrose) anhydrous. The latter contained 2638 mg elemental potassium (green gram 294.6 mg, cow pea 287 mg, coriander seed 247.4 mg, cumin seed 245 mg, oral rehydration salt 1564 mg). The dose was 3 heaped tablespoons, twice a day with meals and sufficient water. The latter provided 1.7-2 gm of potassium. Analysis, safety, and other details of PFS were published,[24].

A five-week supply of pre-packed PFS was provided at baseline and monthly study visits; an unused portion was returned and measured for compliance.

#### Compliance

Participants were adequately counseled to strictly adhere to the allocated interventions and contacted telephonically every 10-14 days to ensure compliance. All participants were aware that the spot urinary assay carried out at monthly intervals will also reflect compliance to the diet.

### Concurrent medication

The previous standard RA medication was continued (background) under rheumatology supervision. Changes in medication were discouraged but allowed if necessary based on clinical judgment. The pain medication was prescribed on a need basis (severe/intolerable pain) and was to be avoided for 12 hours prior to the clinical examination. Medications were carefully recorded. Primary care physicians treated other co-morbid disorders.

### Statistical Plan and Analysis

There was no prior data available to guide the sample size. A modest effect size and a 10% superior response compared to the placebo was assumed for pain relief in Arm B and the sample size was calculated from the tables described in a classic publication,[25]. After adjusting for a 20% dropout rate, 171 subjects were required (80% power, significant p <0.05, two-tailed). There were 57 participants in each study arm.

The diet food record was analyzed by TK in a blinded manner. Standard ‘Food Composition Tables’ (uncooked and cooked foods) were used [22–23]. Allowance was made for potassium and other nutrients in the food supplement. The results of the analysis (diet and food supplement) carried out at week 16 were shown in the current report.

An intention to treat (ITT, last observation carried forward) and per protocol (completer) analysis was carried out. Standard statistical software package (IBMSPSS-20, version 2015 and 2018) was used; parametric (One way ANOVA), non-parametric (Mann Whitney statistic, KW signed rank test), and Chi-square test (categorical data,) and Bonferroni’s correction for repeated measures. Unless stated, all p values in the current report pertain to ANOVA.

Though not intended for the current report, limited results from regression models (Exploratory Analysis, including logistic regression) were shown in Supplement File 1 (Tables 6-8).

## OBSERVATIONS AND RESULTS

One hundred and seventy-two patients were randomized, and 155 (90.1%) patients completed the study (per protocol analysis) (Fig 1). One patient withdrew consent and 171 patients were included in the ITT analysis. Seventeen (9.9%) patients withdrew but not due to an AE.

**Fig 1:**
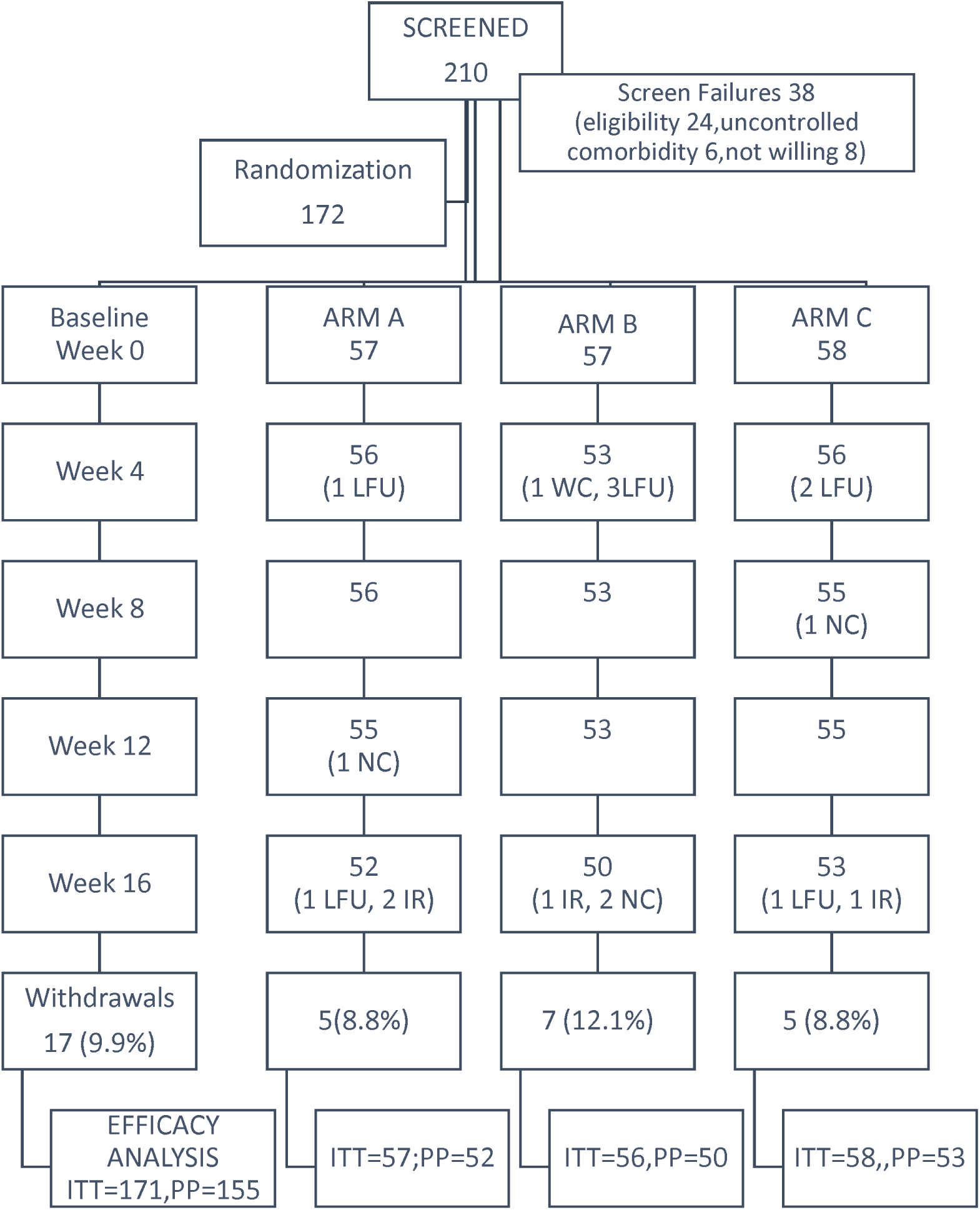
Patient disposition and number withdrawals- A randomized controlled three arm study of Potassium intervention in rheumatoid arthritis (Arm A: potassium rich diet; Arm B: potassium rich diet plus potassium food supplement; Arm C: routine diet; ITT: intention to treat analysis; PP: per protocol analysis; LFU: lost to follow up; WC: withdrew consent; NC: not protocol compliant; IR: inadequate treatment response; see text for details)

At baseline, the study arms were matched(Table 1). Study participants recorded moderately painful active disease (Table 5). Fifty-one patients (29.6%) recorded co-morbid disorders-diabetes (10), hypertension (27), ischemic heart disease (7), chronic acid-peptic disorders (34), hemorrhoids (7), and hypothyroidism (8); the number of patients in parenthesis. None of the participants were found to show any extra-articular complication.

**Table 1:** Baseline demographic and other variables: a randomized controlled potassium diet intervention study in patients suffering from rheumatoid arthritis (RA) and on standard care [A: potassium rich diet; B; potassium rich diet plus potassium food supplement; C; routine diet)

| Variables | A<br>(n=57) | B<br>(n=57) | C<br>(n=58) | P |
| --- | --- | --- | --- | --- |
| Age, mean (years) | 50.5 (11.2) | 47.3 (12.1) | 46.9<br>(10.5) | 0.40* |
| Women (percent) | 85 | 91 | 92 | 0.70** |
| Duration(years)RA, mean, median | 10.6 (8.1), 8 | 8.5 (7.4),6 | 7.4 (5.9), 5 | 0.08* |
| Body Mass Index, mean (Kg/m<sup>2</sup>) | 24.4 (3.8) | 24.3 (4.3) | 24.2(2.9) | 0.90* |
| Pain Score (VAS) > 5 cms (percent) | 52.6 | 54.4 | 50 | 0.87** |
| Erosion hand/joint deformity (percent) | 75.4 | 50.8 | 62.1 | 0.15** |
| Prior continuous DMARD use duration (month), median (range) | 26 (6-240) | 18 (6-360) | 21(6-216) | 0.47* |
| ESR mm fall 1<sup>st</sup> hour, mean | 70.6 (2.2) | 68.8 (3.7) | 67.4 (3.9) | 0.84* |
| C-reactive protein mg/dl, mean | 33.3 (39.4) | 26.2 (31.1) | 25.5 (37.8) | 0.45* |
| RF seropositive (percent) | 76 | 83 | 68 | 0.07** |
| Serum potassium mEq/l, mean | 3.9(0.5) | 3.8 (0.6) | 3.9 (0.5) | 0.59* |
| Urine potassium mEq/l, mean | 44.4 (25.6) | 46.7 (25.8) | 42.9 (29.1) | 0.88* |
| Urine sodium mEq/l, mean | 118.1(79.2) | 109.8 (63.3) | 110 (87.4) | 0.81* |
| DMARD single or combination plus Pred use (percent) | 70.2 | 61.4 | 70.6 | 0.49** |
| DMARD single agent only (percent) | 19.3 | 22.8 | 19 | 0.89** |
| Methotrexate use (percent) | 82.4 | 87.8 | 60 | 0.00** |
| Sulfasalazine use (percent) | 33 | 19 | 47 | 0.01** |
| Hydroxychloroquine use (percent) | 40 | 40 | 47 | 0.27** |
| Analgesic Use^ (percent) | 81 | 95 | 90 | 0.07** |
| Daily Pred dose mg, mean | 5.4 (2.8) | 5.3 (2.5) | 5.5 (2.3) | 0.94* |
| Weekly Methotrexate dose mg, mean | 14.8 (4.4) | 14.9 (4.4) | 13.9 (4.3) | 0.56* |
| Blood Hemoglobin gm/dl, mean | 11.6 (1.3) | 11.6 (1.3) | 12.1 (1.3) | 0.07* |

Note: n: number of patients; Percent pertains to proportion of subjects positive; DMARD: disease modifying antirheumatic drug (methotrexate, sulfasalazine, hydroxychloroquine); Pred: prednisolone; Analgesic ^: includes paracetamol and other analgesic use of non-steroidal anti-inflammatory drug/NSAID (diclofenac, naprosyn, etorocoxib, etodolac, nimesulide) more than 4 times a week : \*Analysis of variance; \*\*Chi-square statistic using count; \*/\*\*; significance: p<0.05, two tailed; mono: single drug; standard deviation shown in parenthesis after mean; see text for details

### Diet

Table 2 shows the analysis at baseline and study completion. Potassium consumption according to RDA is shown in Table 3. The potassium intake at baseline was deficient as per RDA in each of the study arms. Several nutrients increased Arms A and B. The daily potassium intake was substantially increased in Arm B (median 5648 mg, range 4365-7545 mg); 84% of participants consumed 5 gm or more. There were slight/modest changes in Arm C..

**Table 2:** Median (95% confidence interval) of daily energy and nutrient consumption at baseline (week 0) and study completion (week 16) as per protocol analysis: a randomized controlled potassium diet intervention study in patients of rheumatoid arthritis (RA) on standard care [A: potassium rich diet; B; potassium rich diet plus potassium food supplement; C; routine diet)

| Arm/ Nutrient | A<br>(n=52) |  | B<br>( n=50) |  | C<br>( n=53 ) |  | p1 | p2 |
| --- | --- | --- | --- | --- | --- | --- | --- | --- |
| Time points/week | 0 | 16 | 0 | 16 | 0 | 16 |  |  |
| Energy (Kcal) | 2975<br>(2827, 3123) | 3007<br>(2852, 3161) | 2875<br>(2717, 3033) | 3476<br>(3372, 3580) | 2772<br>(2648, 2897) | 2730<br>(2590, 2870) | 0.14 | 0.00 |
| Protein (gm) | 101 (93, 109) | 101<br>(93,111) | 96<br>(88, 103) | 126<br>(111,137) | 89<br>(82, 95) | 89<br>(83, 94) | 0.04 | 0.00 |
| Fat (gm) | 72<br>(69,74) | 76<br>(74,77) | 74<br>(70, 78) | 91<br>(70,133) | 76<br>(74,77) | 77<br>(73,81) | 0.35 | 0.09 |
| Zinc (mg) | 13<br>(12,14) | 14<br>(13,16) | 13<br>(12,14) | 18<br>(16,19) | 13<br>(12,13) | 11<br>(10,20) | 0.46 | 0.11 |
| Calcium (mg) | 800<br>(729,872) | 693<br>(617,769) | 696<br>(638,754) | 1196<br>(1178, | 683<br>(626,739) | 607<br>(566, 649) | 0.01 | 0.00 |

| 1214) |  |  |  |  |  |  |  |  |
| --- | --- | --- | --- | --- | --- | --- | --- | --- |
| Phosphate(mg) | 2198<br>(2058,<br>2340) | 2173<br>(2012,<br>2334) | 2126<br>(1990,<br>2263) | 2515<br>(2216,<br>3625) | 1997<br>(1871,<br>2123) | 1969<br>(1857,<br>2082) | 0.08 | 0.00 |
| Vitamin A (µg) | 1181<br>(1021,<br>1342) | 830<br>(692, 982) | 985<br>(844,<br>1128) | 1032<br>(839,<br>1225) | 1040<br>(836,<br>1244) | 869<br>(696,<br>1042) | 0.34 | 0.17 |
| Thiamin (mg) | 3 (3,3) | 3 (3,3) | 3 (3,3) | 4 (3,6) | 3 (3,3) | 3 (3,3) | 0.57 | 0.05 |
| Riboflavin(mg) | 2 (1,2) | 1 (1,2) | 1 (1,2) | 2 (1,2) | 1 (1,1.1) | 2 (1,2) | 0.05 | 0.27 |
| Niacin (mg) | 25<br>(24,27) | 26<br>(25,28) | 25 (23,26) | 35<br>(23,48) | 24<br>(23,26) | 25<br>(24,27) | 0.65 | 0.07 |
| Vitamin C(mg) | 162<br>(147,177) | 163<br>(151,177) | 152<br>(139,165) | 160<br>(149,172) | 172<br>(153,192) | 169<br>(156,182) | 0.15 | 0.62 |
| Iron (mg) | 34<br>(32,37) | 33<br>(32,36) | 31<br>(29,34) | 41<br>(36,44) | 23.3<br>(23,51) | 30<br>(28,32) | 0.66 | 0.00 |
| Folic Acid (µg) | 421<br>(378,464) | 424.5<br>(386,463) | 393<br>(354,431) | 480<br>(417,543) | 373<br>(334,412) | 308<br>(338,397) | 0.21 | 0.00 |
| Sodium (mg) | 2668<br>(2665,<br>2723) | 2653<br>(2644,<br>2670) | 2649<br>(2820,<br>3038) | 4501<br>(4423,<br>4579) | 2654<br>(2646,<br>2684) | 2647<br>(2636,<br>2658) | 0.17 | 0.00 |
| Potassium(mg) | 2444<br>(2445,<br>2879) | 2959<br>(2832,<br>3282) | 2399<br>(2293,<br>2729) | 6063<br>(5579,<br>6097) | 2469<br>(2324,<br>2858) | 2553,<br>(2428,<br>2920) | 0.65 | 0.00 |

Note: Measure Value rounded to nearest integer and Standard deviation to 1 decimal place; n: number of study participants; p1: p value, ANOVA, comparing baseline (0 weeks); p2: p value, ANOVA, comparing study completion (16 weeks); Food Composition Tables as recommended by National Institution of Nutrition, Hyderabad (See reference serial 23 in text); significant p<0.05, two tailed (ANOVA)

**Table 3:** Median intake and number (percent) of patients achieving recommended daily allowance (India) of potassium and increase intake compared to randomization baseline: A controlled dietary potassium intervention study (n=172) in chronic Rheumatoid Arthritis [A= potassium rich diet; B=potassium rich diet plus potassium food supplement; C= control routine diet]-per protocol analysis

| Arm/Time point<br>week/ Daily<br>Potassium<br>Intake (Target) | A |  | B |  | C |  | P1 | P2 |
| --- | --- | --- | --- | --- | --- | --- | --- | --- |
|  | 0 (n=57) | 16 (n=52) | 0 (n=57) | 16 (n=50) | 0 (n=58) | 16 (n=53) |  |  |
| Daily potassium<br>intake- median<br>(range),90<sup>th</sup><br>percentile , mg | 2902<br>(937-4697),<br>3750 | 2984<br>(713-4619),<br>4150 | 1863.7<br>(939-4162),<br>3400 | 5708.5<br>(4365-7545),<br>6560 | 2697.5<br>(680-4645),<br>3900 | 1749.8<br>(1039-4579),<br>3820 | 0.68 | 0.00<br>(0.02) |
| Minimal RDA<br>(3500 mg)<br>consumption | 14 (25%) | 24 (46) | 12 (21%) | 50(100) | 19 (33%) | 16 (30) | 0.31 | 0.00<br>(0.23) |
| Consumption<br>more than<br>baseline | - | 38 (73) | - | 50 (100) | - | 29 (55) | - | 0.00<br>(0.04) |
| Consumption<br>more than<br>baseline by 20% | - | 23 (44) | - | 48 (96) | - | 15 (28) | - | 0.00<br>(0.10) |
| Consumption<br>more than<br>baseline by 50% | - | 7 (13) | - | 47 (94) | - | 10 (19) | - | 0.00<br>(0.29) |
| Consumption<br>more than 1.5<br>times RDA | - | 0 | - | 42 (84) | - | 0 | - | 0.00 |
| (5250 mg) |  |  |  |  |  |  |  |  |
| Note: Note: n: number of study participants; Target for Arm A: 3200 mg daily (modified RDA for men and women); Target for Arm B: 4800 mg (1.5 times the RDA for women at 3200 mg); 90<sup>th</sup> percentile is rounded to multiple of 50; ,significant p <0.05, two tailed; P1: comparing baseline-ANOVA (median)/Chi-square (categorical data); P2: comparing study completion week 16 -ANOVA (median)/Chi-square (categorical data); parenthesis in P2 shows comparison of Arm A and Arm C (not adjusted for repeated measure); See Text for details |  |  |  |  |  |  |  |  |

Less than 5% of patients (especially in Arm C) recorded consumption of eggs, red meat, fish, or other Western food such as pizza or French Fries. None consumed aerated or alcoholic drinks.

### Serum and Urine Potassium and Sodium Assay (Supplement File 1, Table 1)

All participants remained normokalemic with minor changes(Supplementary File 1, Table 1). The serum potassium was increased, albeit modestly, only in Arm B (0.08, −0.10 0 .24) at week 16. The urine potassium increased significantly in each of the study arm. The mean change at week 16 from baseline was 15.9 (2.3, 28.1) in Arm B, 20.9 (12.3, 33.7) in Arm A, and 16.8 (7.1, 27.2) in Arm C with no significant difference by study arms. Serum sodium remained within normal range at all study time points. However, there was a mild increase (median 5.48) in urinary sodium in Arm B at week 16 from baseline (−16.5, 27.3); corresponding reduction in Arms A and C. Values are mEq/l with 95% confidence interval in parenthesis.

There were no significant difference between the study arms for urinary sodium-potassium ratio at baseline or study completion; at week 16 it was 1.94 (1.31) in Arm A, 2.07 (1.35) in Arm B, and 2 (1.36) in Arm C, standard deviation in parenthesis

### Safety and Tolerability

AE was reported by 14, 16 and 11 participants in Arm A, Arm B and Arm C respectively (p=0.67, Chi-square statistic) (Table 4). There were no serious AE. Mostly they were mild and did not require any medication. Three patients in Arm B reported moderate upper abdominal pain/discomfort which subsided after reducing the dose of the supplement. Laboratory investigations (routine hematology, metabolic renal, and hepatic) as per protocol remained within normal range. Electrocardiography for all participants was reported normal on baseline and study completion.

**Table 4:** Adverse events (number): a randomized controlled potassium diet intervention study in patients suffering from rheumatoid arthritis (RA) and on standard conventional DMARD with/without steroid treatment [A: potassium rich diet; B; potassium rich diet plus potassium food supplement; C; routine diet)

| Adverse Event | A<br>(N = 57) |  | B<br>(N = 57) |  | C<br>(N = 58) |  |
| --- | --- | --- | --- | --- | --- | --- |
|  | Total episode | Number of patients | Total episode | Number of patients | Total episode | Number of patients |
| Nausea | 5 | 4 | 3 | 3 | 4 | 3 |
| Vomiting | 3 | 3 | 1 | 1 | 1 | 1 |
| Acid-peptic symptoms | 3 | 3 | 4 | 4 | 4 | 4 |
| Diarrhea | 1 | 1 | 2 | 2 | 1 | 1 |
| Constipation | 1 | 1 | 3 | 2 | 2 | 2 |
| Anorexia | 2 | 2 | 2 | 1 | 2 | 2 |
| Skin rash | 4 | 4 | 5 | 4 | 2 | 1 |
| Infections | 1 | 1 | 3 | 3 | 1 | 1 |
| Vertigo | 2 | 1 | 0 | 0 | 0 | 0 |
| Oral ulcer | 1 | 1 | 1 | 1 | 0 | 1 |
| Hair fall | 1 | 1 | 0 | 0 | 2 | 1 |
| Leucopenia | 1 | 1 | 0 | 0 | 0 | 0 |

Note : n: number of patients; Infections include upper/lower respiratory tract infections and urinary infections

### Efficacy

Primary Efficacy: Pain (VAS) was reduced in each of the study arms, albeit maximal in Arm B (Fig 2). The mean change at week 16 from baseline was significantly different between the study arms as per protocol analysis (p=0.039) (Table 5). However, the latter was not significant in an intention to treat analysis (p=0.17) (Supplement File 1, Table 2). The mean change in Arm B (95 % confidence interval −2.99, −1.48) was significantly superior to that in Arm C (adjusted p=0.02) and Arm A (adjusted p=0.04) (Table 4); Arm A was superior to Arm C.

**Fig 2:**
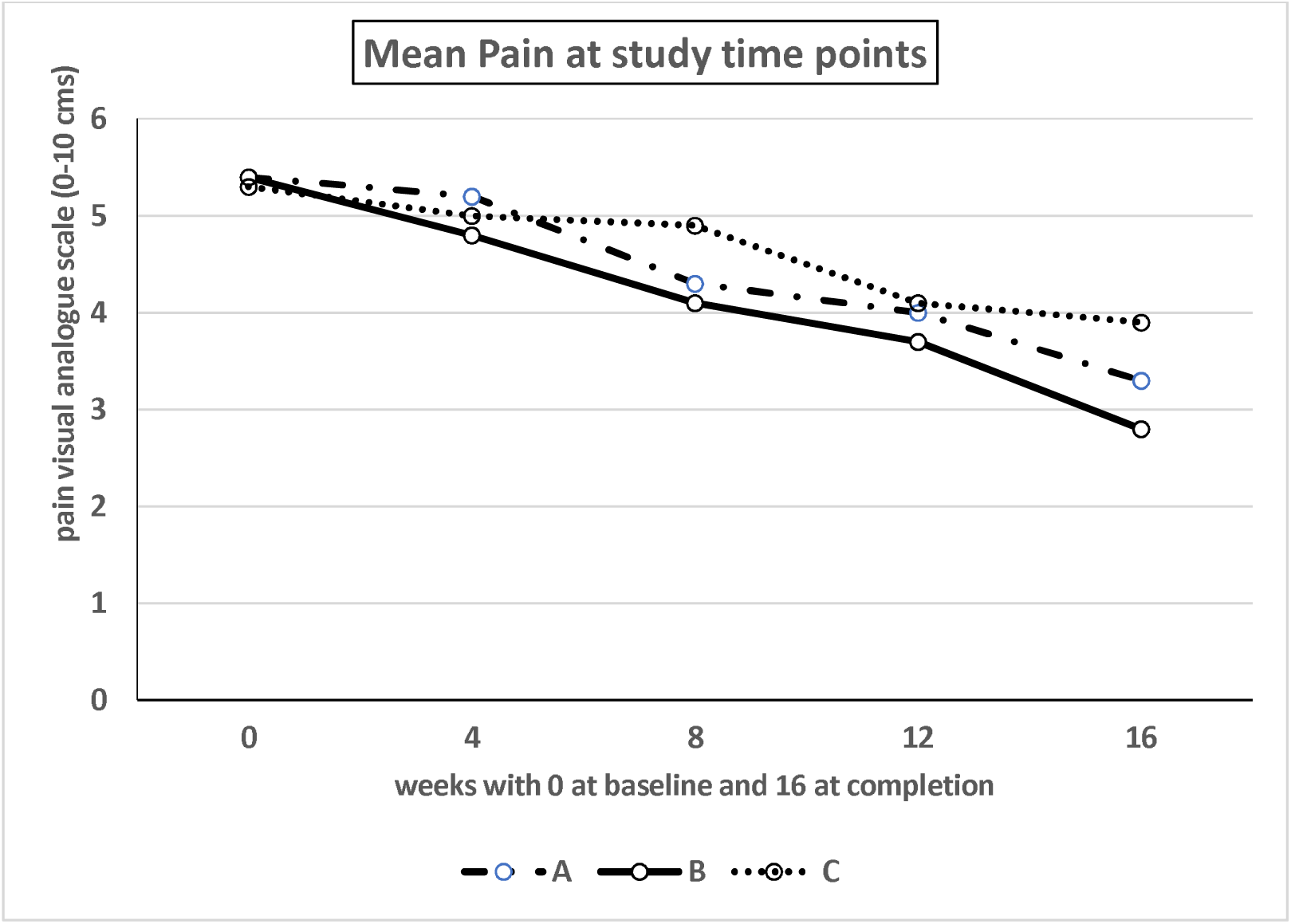
Mean pain visual analogue scale at pre-determine study time points- A randomized controlled three arm study of Potassium intervention in rheumatoid arthritis over 16 weeks duration (Arm A: potassium rich diet; Arm B: potassium rich diet plus potassium food supplement; Arm C: routine diet; see text for details)

**Table 5:**
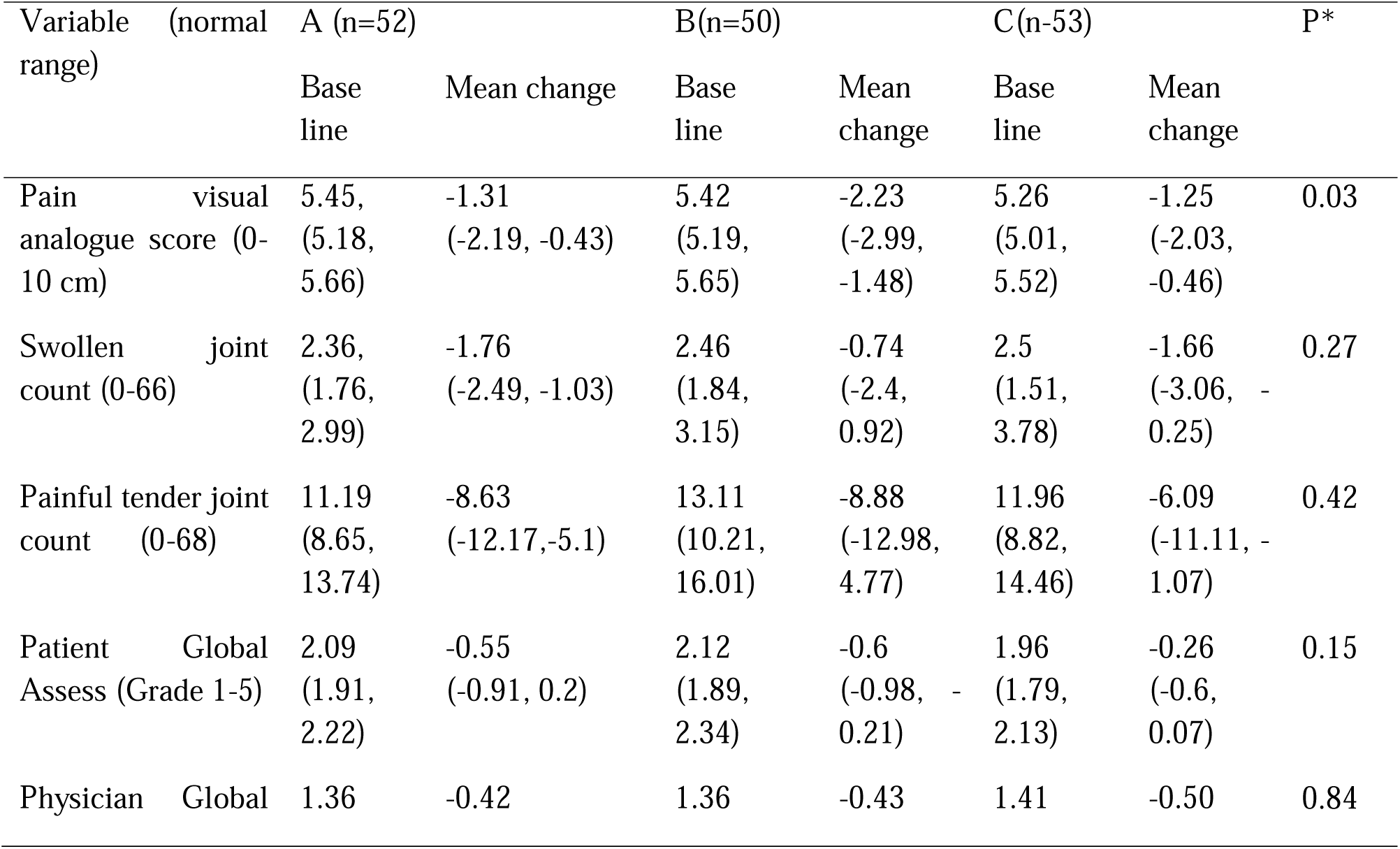
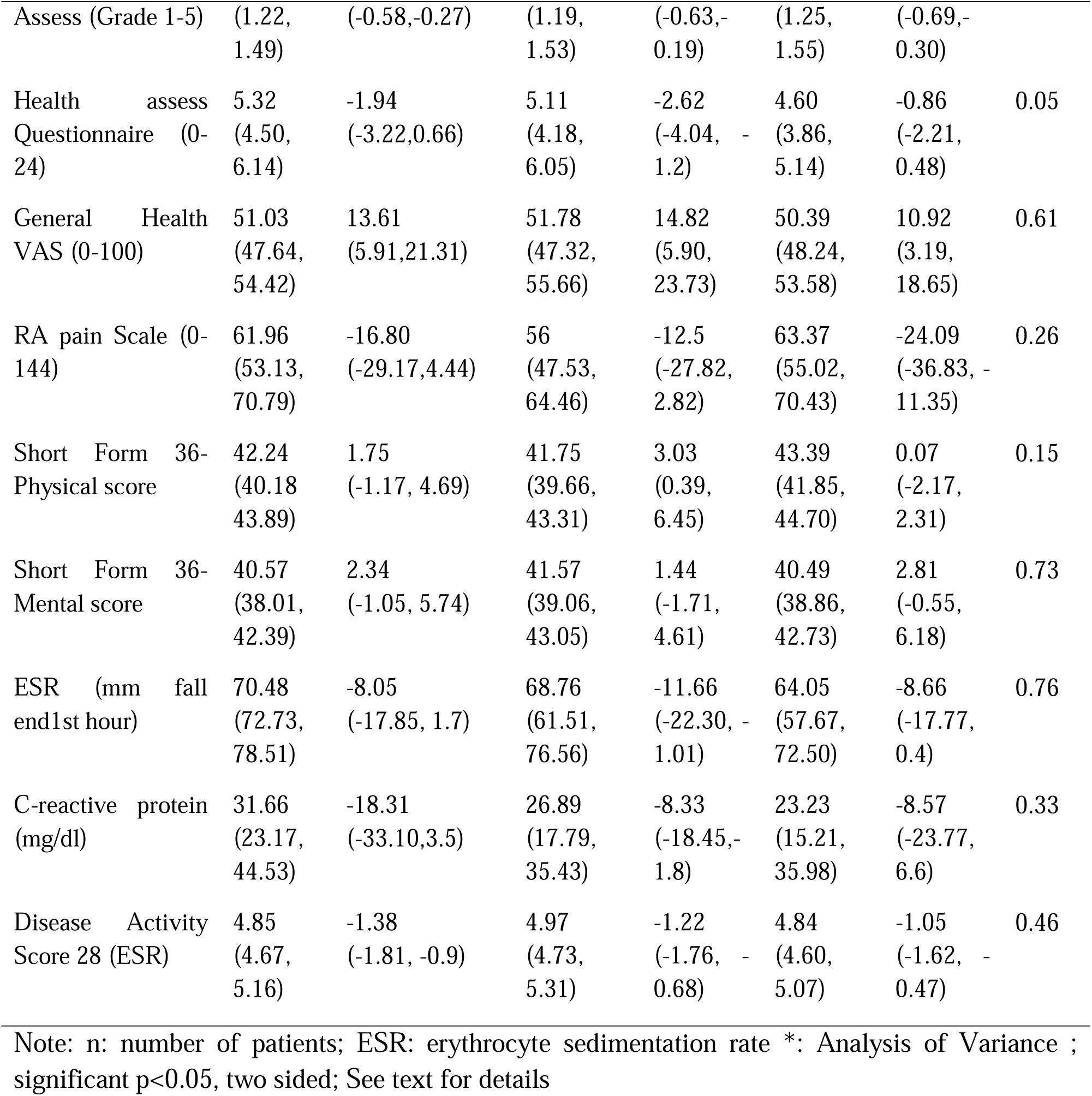
Efficacy Analysis (per protocol)- showing mean at randomization baseline and mean change at week 16 along with 95% confidence interval(in parenthesis): a randomized controlled potassium diet intervention study in patients (n=172) of rheumatoid arthritis (RA)on standard care [A: potassium rich diet; B; potassium rich diet plus potassium food supplement; C; routine diet)

**Table 6:**
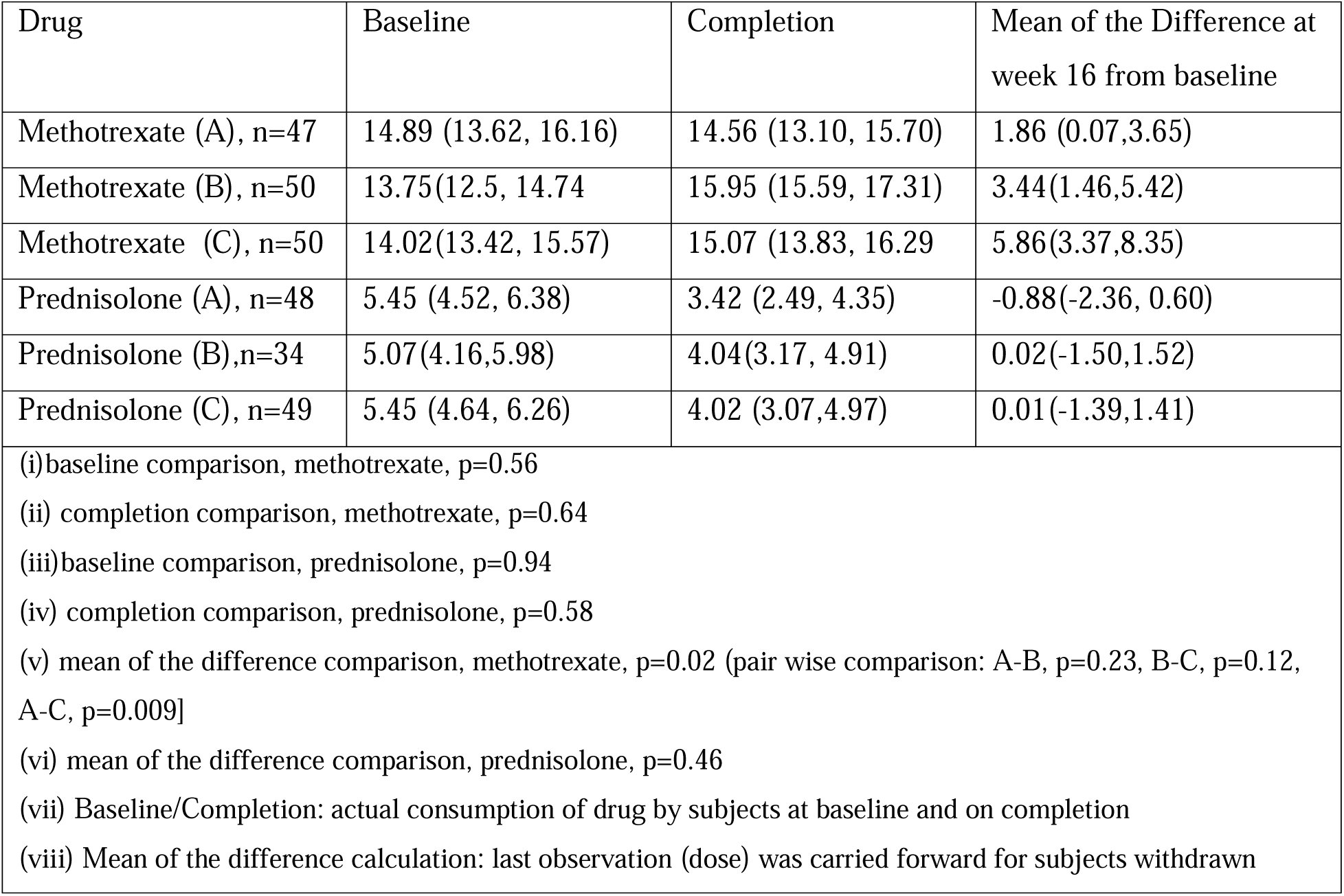
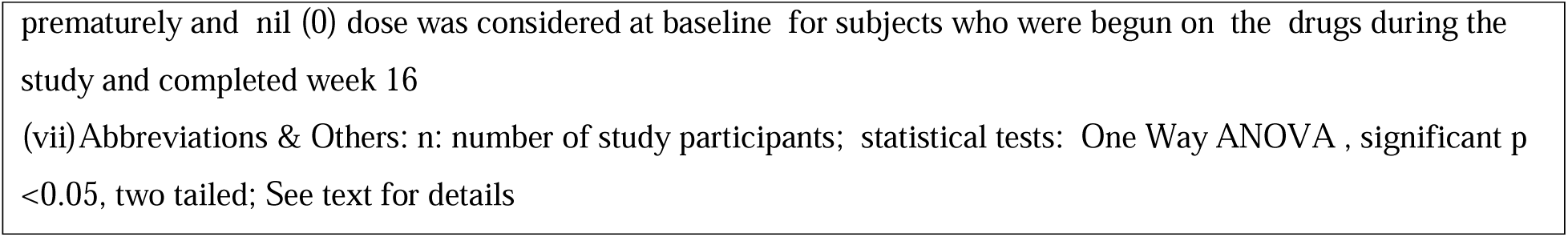
Mean dose (95% confidence interval) of weekly methotrexate (mg) and oral daily prednisolone (mg) at randomization baseline and study completion (week 16) and difference in the mean: A randomized controlled potassium intervention diet study (n=172) in Rheumatoid Arthritis on standard care [A= potassium rich diet; B=potassium rich diet plus potassium food supplement; C= control routine diet]

Secondary Efficacy: Table 5 shows improvement in several variables in each of the study arms. Numerically, maximum number of measures improved in Arm B, and the improvement in joint function (HAQ) and physical quality of life (SF-36) was impressive. The improvement in RA (ACR 20) and disease activity (DAS 28) was modest in each arm and not significantly different. ACR 70 improvement (considered substantial) was only seen in Arm B (5 participants) and Arm A (2 participants).

### RA Medication

The mean doses of weekly methotrexate and daily steroid (low dose) remained stable at baseline and study completion without any significant difference between study arms (Table 6). However, more patients at week 16 in Arm C required methotrexate and there was no significant difference in the use of analgesics (Supplement File 1, Table 4).

### Other Results

Blood pressure: At baseline, the mean systolic/diastolic blood pressure (mm Hg) was 131/79 in Arm A, 129/80 in Arm B, and 125/79 in Arm C; correspondingly at week 16, it was 117/80, 116/80 and 126/82. It was recorded as a routine procedure in a sitting position using a mercury manometer.

Serum Cortisol (morning) assay (Supplementary File 1-Table 1): The maximum increase was observed in Arm B (not significantly different).

Correlation (Supplementary File 1, Table 2, and Table 5): There was a modest inverse correlation between dietary potassium intake and pain VAS (r = −0.19, p<0.05). Urine potassium assay did not correlate with oral potassium intake or serum potassium. There was an inverse correlation between diet potassium and serum potassium both at baseline (−0.345, p<0.01) and completion (−0.129, not significant) (Supplement File 1, Table 2). Diet potassium showed significant correlations with daily energy consumption and several other nutrients (Supplement File 1, Table 3).

Univariate Analysis (Supplementary File 1-Table 7: Daily potassium intake of 5 gm or more (Odds Ratio 3.14) was significantly associated with low pain VAS (≤ 4 cm). Higher urine sodium and potassium excretion individually were significantly associated with low pain VAS. There were no significant associations for DAS 28.

Logistic (Step forward) Regression (Supplementary File 1, Table 8): A daily potassium intake of 5 gm or more (Likelihood Ratio 2.87) and methotrexate use (Likelihood Ratio 16.1) were significant predictors of low pain VAS when adjusted for several clinical, medication and diet-related variables; not significant for any other nutrient.

## DISCUSSION

Participants suffering from symptomatic chronic RA and on background medication showed significant improvement in pain after consuming a potassium-rich diet along with a novel potassium food supplement (Arm B) over the study period. This was a randomized, assessor blind, controlled, single center study of 16 weeks duration. The specific target in the study was to increase the oral potassium intake in active arms (A and B). The latter was considerably exceeded, according to the Indian RDA, in Arm B and considered therapeutic (Table 2). The improvement in pain in Arm A (PRVD), albeit lesser than Arm A, was superior to Arm C (control, routine diet). Though the improvement in disease activity was modest (not significant), it was numerically superior and impressive for joint function and quality of life in Arm B (Table 5)

Importantly, the adverse events were predominantly mild and did not lead to a withdrawal (patient). The compliance (intervention) was satisfactory. The PFS was formulated and locally manufactured under direct supervision by TK. Overall, the safety and tolerability profile of high potassium supplementation was good.

Vegetables, fruits, nuts, cereals, pulses, and dairy products are good resources of potassium,[7, 9]. Potassium may be a marker of a good quality vegetarian diet,[26]. Intracellular shifts, renal mechanism and buffering by muscles maintain body potassium within a narrow plasma range,[9,27]. Potassium and sodium are closely linked in several cellular processes and renal excretion. Dietary intake modulates the activity of Kir 1.4 (potassium channel) and sodium chloride Co-transporter (NCC) in the distal nephron. NCC is pivotal for urinary potassium excretion and this is primarily responsible for the salubrious effect of potassium on blood pressure,[9,27]. An increase in serum potassium stimulates aldosterone which facilitated potassium excretion,[9]. Other mechanisms for regulating potassium excretion include tissue kallikrein (tubular cells) and ion channels (ROMK) [9]. The data on chronic adaptive response to prolonged potassium intake is mostly derived from animal studies [27].

### Strengths and Implication

The study was carried out in a real-life community setting and reflected our clinical practice. The current potassium-driven intervention was encouraged by our earlier experience [11]. Safety and ethical concerns compelled us to execute a diet-based approach for potassium intervention and demonstrate an adjuvant effect. The study was based on a Phase 3 drug trial design. A large sample size (80% power, alpha < 0.05) was enrolled. Ninety percent of patients completed the study. The arms were matched for several variables (Table 1). The latter reduced the bias of potential confounders such as medication and diet and nutrient consumption. There were no safety or tolerability concerns.

Pain is a cardinal RA symptom. Persistent and residual pain is a nuisance despite supervised standard care and was also shown with the use of modern biologic DMARD,[29]. Pain is multifaceted and causes profound deleterious consequences,[1, 18, 29]. The further and superior reduction in pain in both the study Arms A and B as compared to Arm C (control) was indeed encouraging and has considerable implications for the role of diet and potassium in RA. Notably, the mean change in pain VAS in Arm B was within the reported range of minimally clinically important difference (19-27 mm),[28]. In addition, PRVD plus PFS showed improved functionality and quality of life.

There are several complications and other co-morbidity in RA which are likely to benefit from an appropriate diet and potassium. Plant-based diets and potassium supplements are allegedly useful in the management of hypertension and cardiovascular disorders, osteoporosis, dysbiosis and metabolic disorders, sarcopenia, and chronic pain and inflammation,[30–37]. The outcome of the current study encourages research in this direction. Potassium in food supplements is tightly regulated in Europe (500 −1000 mg daily) and the USA (< 100 mg) and some of the guidelines seem outdated,[38,39]. The current study prompts research and revision of the latter.

### Limitations

Several potentially diet-related confounders may complicate the precise interpretation of the study outcome,[40]. We were aware that despite due diligence and care, the latter needed to be addressed at every stage of the current study, beginning with the plan, design and preparing the protocol. We did experience and record several difficulties including those experienced by some participants. Complete adherence to any diet is vexing. Some patients in Arm A did not achieve the RDA for potassium on study completion. Also, it seems that the control diet was influenced to some extent by sharing of information; increase in potassium and some nutrients(Table 2). The food supplement used in the current study was not available in the market. Retrospective recall (diet) can be uncomfortable and inaccurate. Though popularly used as was done in the current study, food composition tables can underestimate the diet content,[41]. Despite the assessor being blind, a placebo response cannot be excluded,[40]. Diet is endearing. Participants in the current study were aware of the dietary nature of the intervention and possibly expected improvement.

Besides potassium, several nutrients with potential health benefits were found increased on study completion (Table 2). Fiber, magnesium, and unsaturated fatty acids were not measured. Plant-based diets were reported to benefit RA,[42]. We have presented data in the current report to support the predominant role of potassium but admit that the vegetarian nature of the study interventions also contributed to the outcome of the current study and that individual effect cannot be dissected out. Notably, higher potassium intake (and not other nutrients) was identified as a significant predictor of low pain in the regression analysis (Supplement File 1, Table 8).

Subtle changes in the background RA medication may have introduced unmeasurable bias in the current study. There may be errors in medicine logs. In our experience, despite substantial pain, patients avoid pain medication because of fear of side effects.

### Other Limitation-Interpretation of Serum and Urinary Assay

Diet potassium intake was associated with urinary potassium assay, lower urinary sodium-potassium ratio, and lower blood pressure in several studies of hypertension including a few in RA,[32, 43,44, 45]. It is prudent to state that the correlation between urine potassium (spot or 24 hours urine) and diet potassium was at the best modest (<0.25) in the latter studies and several more in the published literature. The studies (mentioned here) differed in the methods of diet estimate and analysis, and the urinary assays were invariably cross-sectional. Spot and 24-hour urine samples show high correlation; spot urine assay was a validated method,[32, 43, 46]. The sample size of the RA studies was small and there were selection and other biases,[32, 45].

Several of the current potassium and sodium assay results seem to be at variance with the published literature,[9, 27, 32, 43, 44,45,46]. The expected dosing effect of potassium supplementation in Arm B as compared to Arms A and C was not obvious (Supplement File, Table 2). We found an inverse relationship between diet potassium and serum potassium (see results) which may be due to homeostasis changes. Dietary potassium did not correlate with the spot urinary potassium assay result. In sharp contrast to expectations, the change in urinary potassium assay on study completion was least in Arm B (potassium supplementation). We do not have a plausible explanation for the latter but this may be due to chronic adaptation and unique factors in RA (see below). Serum and urine sodium remained unchanged though the sodium intake was considerably increased in Arm B (Table 2). Participants did not follow any individual schedule and there may be some error in urine sample collection and assay. Renal potassium secretion is optimum by mid-day /noon (circadian rhythm),[9]. A systematic bias was unlikely.

The current study diet was also rich in carbohydrate which is known to stimulate insulin secretion and in turn facilitate intracellular shift (especially muscles) of potassium,[9]. There may be another caveat. To begin with, the baseline diet of the participants seemed inadequate for potassium (Table 2). And this was our earlier experience also,[11]. Perhaps, the participants had total body deficiency for potassium which we did not estimate. Interestingly, a study from the UK reported low body potassium in chronic RA based on isotope studies,[47].

The pathophysiology of potassium in chronic RA is likely to be complicated by several other factors -chronicity of immune-mediated inflammation, systemic nature of the disease, prolonged intake of drugs especially steroids and non-steroidal anti-inflammatory drugs, muscle wasting (sarcopenia), and frequent co-existent morbidity (in particular cardiovascular and endocrine).

### Mechanism of Action

We do not know the precise mechanism for the reduction in RA pain by diet and supplement-based intervention in the current study. We focus on potassium. The reduction in pain may be due to a non-specific anti-inflammatory effect and this may be partly mediated by increased endogenous steroids. This was also speculated earlier,[10,12]. In the current study, though not statistically significant, the morning serum steroid assay was maximally increased in Arm B (Supplement File-Table 1).

Modulation of potassium ion channels reduced pain and inflammation in chronic pain states and RA, [8, 36,37]. These cell surface channels are ubiquitous and also regulate and modulate the nerve impulse (action potential) for pain. Several potassium channels operate in the renal collecting duct system to regulate potassium. However, research is warranted on potassium channels in RA.

Higher salt intake, and sodium in particular. may facilitate immune-mediated inflammation in RA, [48]. It is plausible that increased potassium intake indirectly ameliorates the sodium-induced effect.

### Other Food and Diet Intervention Studies in Chronic RA

The American College of Rheumatology(ACR) recently recommended an integrative approach and advocated a Mediterranean-style diet (MD) in the management of RA[49].

A recent comprehensive meta-analysis of several MD intervention studies in RA endorsed a a modest anti-inflammatory effect[50]. A meta-analysis of 7 randomized controlled trials (RCT) using anti-inflammatory diet, mostly MD, showed a significant, albeit modest, reduction in pain mean change −9.22mm/100 mm VAS, 95% CI −14.15 to −4.29),51]. Sample sizes were small and inadequate (low statistical power), and there were several biases (especially patient selection and RA status).

Further significant improvement in several RA measures was shown by addition of MD to chronic RA on supervised prolonged standard care in each of the 2 RCT (control routine diet); one (Swedish study) at week 12 and the other (UK study) at week 24,[52,53]. A recent RCT (ADIRA) of 10 weeks duration failed to show an advantage of MD (plus probiotics) over routine Swedish diet to further manage patients of chronic RA (on prolonged standard treatment) [54].

There are very few long-term diet interventional drug trials. Significant reduction in pain (mean change −1.89/10 cm VAS, 95% confidence interval −3.62, −0.16) with addition of a vegetarian diet on study completion in a RCT of 13 months in symptomatic chronic RA (on treatment) as compared to routine diet [55]. A similar improvement was shown by addition of a gluten-free vegan diet to patients of chronic RA (on treatment) as compared to balanced vegetarian diet in a one year RCT; antibodies to gliadin and lactoglobulin were reduced [56].

Two recent plant-based diet (PBD) interventional RCT studies in chronic RA on stable medication showed modest reduction in arthritis (DAS 28) [57,58].Life style measures were advocated with PBD in the Dutch study of 16-week duration; also showed metabolic benefits as compared to routine diet [57]. In the German study of 12 weeks duration PBD with an emphasis on turmeric, cinnamon, and coriander was as compared to a control routine anti-inflammatory diet [58].

The data on potassium in MD and PBD trials is sparse. Potassium loaded with antioxidant vitamins, fibre, and flavonoids, as the first pattern in a principal component analysis to identify dietary patterns to explain the variation in reduced arthritis activity in patients of chronic RA on standard drugs who routinely consumed a MD like diet [59]. A higher adherence to MD was associated with an increased potassium intake (17% meat, 15% vegetable, 15% fruit) in a recent Italian community study [60].

The characteristics of MD are well known-plenty of vegetables and fruits, unrefined cereals, olive oil, and moderate intake of dairy products; use of herbs and spices are encouraged while red meal and sugars are discouraged [49,52]. In contrast, while adhering to the diet intervention allocation in the current study followed traditional Indian cooking and consumed cooked vegetables and pulses using oil and several spices along with cooked rice and Indian bread (wheat or millet); generous use of turmeric, ginger, coriander, green chili, onion, and tomato was customary. Consumption of non-vegetarian food and eggs was negligible.

## Conclusion

Patients of symptomatic chronic RA consuming a daily intake of PRVD with or without a PFS showed significant and substantial improvement in pain and joint function on study completion in randomized, assessor blind, controlled study of 16 weeks duration. Participants were on supervised and stable RA medication. The high potassium-based intervention (5-7.5 gm daily) was found safe and well tolerated and none of the AE were severe. We recommend a potassium-rich predominantly vegetarian diet in all patients of RA. In addition, patients with difficult and chronic RA may benefit from a judicious adjunct use of potassium supplementation. Further research and validation studies in different global settings are required.

## Data Availability

All data produced in the present study are available upon reasonable request to the authors

## ACKNOWLEDGMENT

Arthritis Research Care Foundation-Centre for Rheumatic Diseases CRD), Pune, India, provided generous material and logistic help for the study. Several colleagues (rheumatologist) assisted in the study and namely Dr Nachiket Kulkarni, Dr Naisar Nahar, and Dr Kiran Adams. Several administrative, nursing and paramedic staff of CRD Pune provided wholehearted assistance and participation. We remain indebted to our patients who volunteered and consented to participate in the study despite several personal and logistic hurdles.

## AUTHORS CONTRIBUTION

All the authors actively participated in drafting and finalizing the version of the current manuscript prior to submission.

Dr Toktam Kianifard and Dr Arvind Chopra shared equally to this manuscript as First Author.

Dr Toktam Kianifard and Dr Arvind Chopra had full access to all of the study data and data analysis and take full responsibility for its integrity and accuracy.

All the authors approved the final version of the manuscript to be published. Study Conception and design. Kianifard, Chopra

Acquisition of data. Kianifard, Saluja, Venugopalan

Analysis and Interpretation. Kianifard, Sarmukkaddam, Saluja, Venugopalan, Chopra

## DATA SHARING

The raw data and any other relevant study data duly anonymized for the study participant is available with Dr Arvind Chopra (Senior Author) and access will be provided to an applicant from a non-commercial settings one year after the manuscript is published. The application should be submitted to Dr Arvind Chopra by Email explaining the reason for access, employability details, protocol and funding of the proposed study, contact details and CV.

