## Supplement File 1 for "An Adjunct Role Of Potassium Rich Vegetarian Diet And A Novel Potassium Food Supplement To Improve Pain In Chronic Rheumatoid Arthritis On Supervised Standard Care: A Randomized Controlled Study"

Main Text Title- Adjunct Role of Potassium in Painful Rheumatoid Arthritis: A Randomized Controlled Study of Diet and Food Supplement based Intervention in Patients on Supervised Standard Care

**SUBJECT: Additional Efficacy Results**

Table 1: Median (Standard deviation, 95% confidence intervals) serum cortisol, serum and urine potassium over 16 weeks study period: A randomized controlled potassium-diet study in 172 patients of chronic rheumatoid arthritis on standard treatment[Arm A= potassium rich vegetarian diet, B= potassium rich vegetarian diet plus potassium food supplement, C=control routine diet]-per protocol analysis

| Variable | Gp | Baseline | 4 weeks | 8 weeks | 12 weeks | 16 weeks | p* | P1 | P2 |
| --- | --- | --- | --- | --- | --- | --- | --- | --- | --- |
| Serum K (mEq/L) | A | 3.9 (0.5) | 3.8 (0.4) | 3.9(0.5) | 3.9(0.5)) | 3.6 (0.5) | 0.21 | 0.59 | 0.36 |
|  | B | 3.8(0.5) | 3.8(0.4) | 3.7 (0.4) | 3.9 (0.5) | 3.9 (0.4) | 0.35 |  |  |
|  | C | 3.9 (0.5) | 3.7 (0.4) | 3.7 (0.5) | 4 (0.5) | 3.8(0.5) | 0.99 |  |  |
| Serum Na (mEq/l) | A | 140 (6.6) | 139 (5.1) | 141.5 (5) | 142 (6.7) | 139 (9.3) | 0.52 | 0.67 | 0.41 |
|  | B | 140(5.4) | 137 (4.7) | 141.5 (9.8) | 142 (7.2) | 139 (9.1) | 0.40 |  |  |
|  | C | 142 (6.1) | 139 (4.3) | 139 (6.1) | 143 (8.9) | 140 (8.1) | 0.49 |  |  |
| Urine K (mEq/l) | A | 37.6 (25.9) | 39 (39.1) | 51.5(32.3) | 65(42.1) | 65.5(34.7) | 0.01 | 0.77 | 0.50 |
|  | B | 40 (26.1) | 43 (29.6) | 48 (34.1) | 45 (23.6) | 59 (38.5) | 0.02 |  |  |
|  | C | 34.8 (28.9) | 45 (39.8) | 45 (30.4) | 47 (27.8) | 55 (36.6) | 0.00 |  |  |
| Urine Na (mEq/l) | A | 105.5(79.2) | 81 (59.2) | 95.5 (45.2) | 65(93.3) | 117 (44.7) | 0.51 | 0.81 | 0.82 |
|  | B | 97(63.4) | 94.5(84.4) | 98.5(70.8) | 81(55.3) | 116 (53.8) | 0.61 |  |  |
|  | C | 99(87.4) | 104(66.6) | 100(58.5) | 101(55.4) | 105(43.9) | 0.53 |  |  |
| Serum Cortisol (µg/dl)  (8-1030 am) | A | 5.7 (4.2) | 6.8 (3.9) | 7.6 (7.1) | 8.4 (8.9) | 8.2 (7.8) | 0.00 | 0.59 | 0.55 |
|  | B | 5.5 (4.8) | 6.5 (5.7) | 6.9(6.6) | 9.4 (9) | 8.9(8.5) | 0.00 |  |  |
|  | C | 5.4 (4) | 7.6 (7.6) | 6.3 (6.6) | 8.1 (7.6) | 7.1 (7) | 0.01 |  |  |
| Note: K: potassium; Na: sodium; Gp: group; Number of patients: 52 Arm A, 50 Arm B & 53 Arm C; Number of Serum samples: 100% serum samples at baseline and completion and 90-94% other visits; Number of Spot Urine Potassium samples: least 48 available at each visit; Urine sample collected for spot urine K assay between 8-11 am; Blood collected for serum cortisol between 8-1030 am; Significant p<0.05two-tailedd (ANOVA); No significant differences at p<0.05 between groups at study visits using ANOVA ;p*: in-between group change: p1:baseline comparison ; p2: study completion (week 16) comparison; See main text for details | | | | | | | | | |

Table 2: Correlations (r) between potassium (K) related diet and laboratory variables at baseline and completion and with selected outcomes in patients with symptomatic rheumatoid arthritis (RA) randomized (n=172 patients) to a diet intervention drug trial; data pertains to 155 patient completers.

| Variable | Diet K (B) | Diet K (C) | Serum K (B) | Serum K (C) | Urine K (B) | Urine K (C) |
| --- | --- | --- | --- | --- | --- | --- |
| Diet K (B) | 1 | 0.038 | -0.345** | -0.127 | -0.056 | 0.085 |
| Diet K (C) | 0.038 | 1 | -0.129 | 0.021 | 0.002 | -0.089 |
| Serum K (B) | -0.345** | -0.129 | 1 | 0.304** | 0.086 | -0.09 |
| Serum K (C) | -0.127 | 0.021 | 0.304** | 1 | 0.03 | -0.94 |
| Urine K (B) | -0.056 | 0.002 | 0.086 | 0.03 | 1 | 0.051 |
| Urine K (C) | 0.085 | -0.089 | -0.09 | -0.94 | 0.051 | 1 |
| Pain VAS (C) | -0.009 | -0.193* | -0.081 | -0.068 | 0.114 | 0.115 |
| Pain MCID (C) | 0.114 | -0.191* | -0.071 | -0.105 | 0.094 | 0.088 |
| DAS 28 (C) | 0.018 | 0.006 | -0.041 | 0.029 | -0.008 | 0.079 |
| Note: n:number; B:caseline; C:completion; Diet: daily estimation based on ‘Food Composition Table (India); Urine K: spot morning urine assay; Pain VAS: pain visual analogue scale; MCID: minimum clinically important difference; DAS 28: disease activity score based on 28 joints; *:p<0.05; **:p<0.01; See methods above and main test for further explanation | | | | | | |

Table 3: Efficacy variables of patients suffering from RA on standard of care treatment in intervention and control group (A; K rich diet , B; K rich diet + dietary K suppl C; routine diet): showing mean change (95% confidence interval) over study period (16 weeks) - an Intention to treat analysis.

| Variable | A(n=57) | | B(n=58) | | C(n=57) | | P*  ANOVA |
| --- | --- | --- | --- | --- | --- | --- | --- |
|  | Baseline | Mean  change | Baseline | Mean  change | Baseline | Mean  change |  |
| Pain VAS  (0-100mm) | 5.42 | -1.31  (-1.93,-0.7) | 5.41 | -1.98  (-2.62,-1.34) | 5.26 | -1.24  (-1.8,-0.67) | 0.17 |
| JCSW  (range 0-66) | 2.37 | -1.82  (-2.37,-1.27) | 2.49 | -0.69  (-1.88,0.48) | 2.44 | -1.60  (-2.61,-0.6) | 0.2 |
| JCPT  (range 0-68) | 11.19 | -9.23  (-11.82,-6.64) | 13.11 | -8.58  (-12.11,-5.05) | 11.64 | -5.78  (-9.38-2.19) | 0.27 |
| Patient Assess (Grade 1-5) | 1.35 | -0.42  (-0.58,-0.27) | 1.35 | -0.41  (-0.63,-0.19) | 1.41 | -0.5  (-0.69,-0.3) | 0.21 |
| HAQ  (range 0-24) | 5.32 | -1.89  (-2.85,-0.93) | 5.11 | -2.14  (-3.38,-0.91) | 4.5 | -0.72  (-1.73,0.27) | 0.13 |
| General Health  (0-100 mm VAS) | 51.03 | 13.44  (8.05,18.83) | 51.49 | 13.41  (6.76,20.06) | 50.91 | 10.30  (4.58,16.02) | 0.68 |
| SF36 Physical score | 42.03 | 1.78  (-0.3, 3.87) | 41.48 | 2.83  (0.40, 5.26) | 43.28 | 0.31  (-1.38, 2.01) | 0.23 |
| SF36 Mental score | 40.20 | 2.5673  (-0.01,5.15) | 41.05 | 1.30  (-0.94, 3.55) | 40.80 | 2.29  (-0.43, 5.02) | 0.76 |
| ESR mm fall 1^st^ hour | 70.62 | -8.38  (-15.62, -1.13) | 69.03 | -10.88  (-18.75, -3.02) | 65.08 | -9.29  (-15.95, -2.63) | 0.88 |
| CRP mg/dl | 33.85 | -22.02  (-35.16, -8.87) | 26.61 | -8.33  (-15.90, -0.75) | 25.6 | -11.74  (-23.55, 0.06) | 0.2 |
| DAS 28 ESR | 4.91 | -1.46  (-1.79, -1.14) | 5.01 | -1.18  (-1.60, -0.77) | 4.83 | -1.02  (-1.44, -0.60) | 0.25 |
| Note: n: number of patients; VAS: visual analogue scale; JCSW: swollen joint count; JCPT: painful joint count; HAQ: health assessment questionnaire (function); SF 36 : Short Form 36 (quality of life); ESR: erythrocyte sedimentation rate; CRP:C-Reactive protein; DAS: disease activity index; Higher value/scores at baseline except for general health and SF 36 indicate worst outcome; Normal ranges are shown in parenthesis after variable; See Text for details | | | | | | | |

Table 4: Rheumatoid Arthritis Medication (number of patients) at randomization baseline (week 0) and study completion (week 16) in a randomized controlled three arm dietary potassium intervention study (n=172) in chronic Rheumatoid Arthritis [A= potassium rich diet; B=potassium rich diet plus potassium food supplement; C= control routine diet]

| Arm/ Drug | A | | B | | C | | p1* | p2* |
| --- | --- | --- | --- | --- | --- | --- | --- | --- |
| Time points/week | 0  (n=57) | 16 (n=52) | 0 (n=57) | 16 (n=50) | 0 (n=58) | 16 (n=53) |  |  |
| **DMARD (Single or Combo) + Prednisolone/P (low dose steroid, 5 mg or less daily dose)** | | | | | | | | |
| DMARD (Single or combination) plus prednisolone-total | 40 | 35 | 35 | 32 | 41 | 39 | 0.49 | 0.56 |
| Methotrexate(P) | 16 | 13 | 20 | 17 | 12 | 17 | 0.2 | 0.29 |
| HCQS (P) | 8 | 0 | 2 | 1 | 6 | 2 | 0.12 | 0.46 |
| Sulfasalazine (P) | 0 | 2 | 1 | 0 | 6 | 1 | 0.04 | 0.35 |
| Methotrexate + HCQS (P) | 3 | 8 | 6 | 8 | 5 | 6 | 0.46 | 0.84 |
| Methotrexate + Sulfasalazine (P) | 8 | 8 | 4 | 4 | 5 | 7 | 0.57 | 0.42 |
| Sulfasalazine+ HCQS (P) | 0 | 0 | 0 | 0 | 5 | 1 | 0.08 | 0.37 |
| Methotrexate + Sulfasalazine+ HCQS (P) | 5 | 4 | 2 | 2 | 2 | 5 | 0.34 | 0.35 |
| **DMARD Combination (no prednisolone)** | | | | | | | | |
| DMARD combination-total | 11 | 12 | 13 | 12 | 11 | 10 | 0.89 | 0.41 |
| Methotrexate + Sulfasalazine | 5 | 5 | 3 | 3 | 3 | 5 | 0.76 | 0.98 |
| Methotrexate + HCQS | 5 | 7 | 9 | 8 | 4 | 5 | 0.28 | 0.23 |
| Sulfasalazine+ HCQS | 1 | 0 | 1 | 1 | 4 | 0 | 0.99 | 0.42 |
| **DMARD Mono** | | | | | | | | |
| HCQS | 1 | 0 | 3 | 0 | 1 | 0 | 0.23 | 0.26 |
| Sulfasalazine | 0 | 0 | 0 | 0 | 2 | 0 | - | - |
| Methotrexate | 5 | 5 | 6 | 6 | 3 | 4 | 0.81 | 0.81 |
| **Total Use of DMARD (Single/Combo with or without Prednisolone)** | | | | | | | | |
| Methotrexate | 47 | 50 | 50 | 48 | 34 | 49 | 0.00 | 0.60 |
| Sulfasalazine | 19 | 19 | 11 | 10 | 27 | 19 | 0.01 | 0.14 |
| HCQS | 23 | 19 | 23 | 20 | 27 | 17 | 0.27 | 0.88 |
| **Analgesic/NSAID Use** | | | | | | | | |
| Analgesic/NSAID | 46(81) | 40(75) | 54(95) | 39(75) | 52(90) | 43(81) | 0.07 | 0.93 |
| **Equivalent paracetamol** daily use ^ , gm, mean (SD) | 1.85 (0.82) | 1.37 | 1.8 | 1.41 | 1.9 | 1.46 | 0.83 | 0.85 |
|  |  | -0.7 | -0.77 | -0.77 | -0.85 | -0.7 |  |  |
| Note: n: number of study participants; DMARD: disease modifying antirheumatic drug; Analgesic/NSAID^: Daily Analgesic use in varying and/or fixed dose >4 times a week (includes non-steroidal anti-inflammatory drugs/NSAID) ; Daily paracetamol use^^: Combined use of paracetamol and Non-steroidal anti-inflammatory (NSAID) whereby NSAID use was converted into equivalent paracetamol dose by an equation decided a-priori by expert consensus (Each tablet of 50 Diclofenac/100 mg Nimesulide/60 mg Etorocoxib/300 mg etodolac were equated to 1000 mg paracetamol; *p1: baseline comparison of groups; *p2: completion comparison of groups: *: chi-square statistic (Yates correction), degrees of freedom 2, significant p <0.05; See text for details | | | | | | | | |

Table 5: Significant Pearson Moment correlation (r) between Diet nutrients and Energy Consumption at Baseline (B) and Study Completion (C) in patients with symptomatic RA randomized (n=172 patients) to a diet intervention drug trial; Data pertains to 155 patient completers.

| **Diet Variable** | **Positive ‘r’** | **Negative ‘r’** |
| --- | --- | --- |
| Potassium (B) | Sodium , Iron (C), | Protein(C),Zinc(C),Calcium (C),Thiamine (C) , Folic A(C) |
| Energy (B) | Protein (B)*, Zinc (B)*,Calcium (B), Thiamine(B)*, Iron (B), Folic acid (B) **, Sodium (B), Protein (C ), Zinc(C ), Calcium (C ), Thiamine (C )*, Iron (C) *, Folic acid (C)* |  |
| Protein (B) | Energy (B) *, zinc (B)*, calcium(B)*, thiamine (B)*, iron (B), folic acid (B) **, sodium (B), protein (C ), zinc(C ), calcium (C ), thiamine (C)*, iron (C) * | Nil |
| Fat (B) | Nil | Nil |
| Calcium (B) | Energy (B), Protein (B)*, Zinc (B), Vitamin A (B), Thiamine (B) *, Iron (B)*, Folic acid (B) *, Sodium (B)*, Protein (C ), Thiamine (C) , Iron (C ) | Nil |
| Thiamine (B) | Energy (B) *, Protein (B)*, Zinc (B)*,Calcium (B)*, Iron (B)*, Folic acid (B) *, Sodium (B), Protein (C )*, Zinc (C ), Calcium (C ), Thiamine (C )*, Iron (C ) *, Folic acid (C )* | Nil |
| Vitamin C (B) | Nil | Calcium (C ), Thiamine (C ), Iron (C), Folic acid (C ) |
| Iron (B) | Energy (B), Protein (B), Zinc (B), Calcium (B), Vitamin A (B), Thiamine (B) *, Folic acid (B), Sodium (B) | Nil |
| Folic Acid (B) | Energy (B)*, Protein (B)*, Zinc (B)*, Calcium (B), Vitamin A (B), Thiamine (B)*, Iron (B)*, Sodium (B), Protein (C )*, Zinc (C ), Calcium (C) , Thiamine (C )*, Iron (C ) | Nil |
| Vitamin A (B) | Calcium (B)*, Iron (B), Folic acid (B) | Nil |
| Zinc (B) | Protein (B)*, Thiamine (B)*, Iron (B), Calcium (B), Folic acid (B) *, Sodium (B), Protein (C ), Thiamine (C )*, Iron (C ), folic acid (C ) | Nil |
| Sodium(C) | Energy (C)*, Protein (C)*, Fat (C), Zinc(C)*, Calcium (C)*, Phosphate(C), Thiamine (C)*, *Iron (C)*, Folic acid (C)*, Vitamin A(C), Potassium (C) | Nil |
| Potassium (C) | Energy (C), Protein (C)*, Fat (C), Zinc (C)*, Calcium (C ), Phosphate(C), Thiamine (C), Iron (C ), Folic acid (C), Sodium (C)* | Vit C (B) |
| Note: n: number ; Diet variables: measured as daily quantity based on standard ‘Food Composition Tables (India)’; Abbreviations and acronyms: see above methods; Significance at p<0.05 two tailed; * : p<0.01; See main text for further details | | |

Table 6: Variables used in Univariate and Logistic Regression Analysis: Definition and Classification of Variables (dummy binary codes- 1 and 2) and Dependent Variables- A Randomized Assessor Blind three Arm Controlled Diet Intervention Study of Symptomatic Rheumatoid Arthritis (RA) (n=172 patients) of 16 Weeks Duration.

| **Variables for Regression Analysis** | **Variable Label** | **Explanatory note / dummy code 1 (equivalent to Yes)** |
| --- | --- | --- |
| Potassium /K_Arm | K_Arm | Arm A or B, consumed potassium |
| K diet arm | K_5_diet arm | Arm A only |
| Age continuous (years) | Age | continuous data |
| Age stratified (years) | Age_40l | age less than 40 years |
| Duration (years) | R_5m | 5 years or more |
| Tobacco | Tobacco | Yes |
| Menopause | Menopause | Yes |
| BMI stratified | BMI_25m | 25Kg/m^2^ and more=overweight and obesity |
| Joint count pain tender (JCPT) | JCPT_1_7m | 7 joints or more painful or tender baseline |
| Joint count swelling (JCSW) | JCSW_1_2m | 2 joints or more swollen baseline |
| Health Assessment Questionnaire (HAQ) | HAQ_1_6m | 6 (total 24) or more disability score baseline, more disability |
| Physician global assess | PGA_1_3m | 3 or more category physician global assess disease severity baseline |
| Patient assessment disease (PAD) | PAD_!_3m | 3 or more category patient global assess disease severity baseline |
| General Health Assess (GHA) | GHA_1_60l | 60mm (VAS) or less score baseline to show more poor health |
| Early Morning Stiffness (EMS) | EMS_1_30m | 30 min or more morning stiff baseline, more severe disease |
| Rheumatoid Arthritis Pain Score (RAPS) | RAPS_1_60m | 60 or more baseline score for more pain |
| Disease activity score (DAS) | DAS_!_5.1m | DAS28 high on baseline > 5.1 more disease active |
| Short Form Health Score- physical 36 item (SF36P) | SF36P_1_40l | 40 or less score baseline for more physical disability |
| Short Form Health Score- mental 36 item (SF36M) | SF36M_1_40l | 40 or less score baseline for more mental disability |
| Erythrocyte Sediment Rate (ESR) | ESR_1_50m | 50mm fall 1^st^ hour or more measure baseline for more disease severity |
| Serum Potassium (Sr K) | SrK_1_3.5l | 3.5mEq/L or less assay baseline for lesser body potassium |
| Urine Potassium (K) | UrK_1_40m | 40mg or more excretion baseline for more K loss |
| C-reactive protein (CRP) | CRP_1_12m | 12mg/dl or more assay baseline for more disease severity |
| Rheumatoid Factor (RF) titre | RF_1_120m | 120 IU/l or more assay baseline for more seropositive RA |
| Anti-cyclic citrullinated peptide (CCP) assay | CCP_1 | >5 RU/l |
| Serum Cortisol (Sr Cort) | SrCort_1_7.5 less | Serum cortisol less than 7.5 mg baseline in more painful diseases |
| Serum Cortisol (Sr Cort) | SrCort_5_7.5 more | Serum cortisol more than 7.5 mg completion in less painful diseases |
| Energy- daily diet consumption | Energy_1_2700m | 2700 KCalories or more baseline Kcal consumption |
| Protein- daily diet consumption | Prot_1_80m | 80gm or more protein baseline consumption |
| Zinc-daily diet consumption | Zn_1_15m | 15mg or more zinc baseline consumption |
| Vitamin C (Vit) daily consumption | VitC_1_130m | 130mg or more Vit C baseline consumption |
| Iron -daily diet consumption | Iron_1_30m | 30mg or more Iron baseline consumption |
| Potassium (K)-daily diet consumption | K_1_2200l | 2200 mg or less baseline consumption |
| Energy- daily diet consumption | Energy_5_2700m | 2700 KCaolories or more Kcal consumption on study completion |
| Protein- daily diet consumption | Prot_5_80m | 80gm or more protein consumption on study completion |
| Zinc-daily diet consumption | Zn_5_15m | 15mg or more zinc consumption on study completion |
| Calcium consumption diet | Cal_5_800m | 800 mg or higher calcium consumption study completion |
| Vitamin C (Vit) daily consumption | VitC_5_130m | 130mg or more Vit C consumption on study completion |
| Iron -daily diet consumption | iron_5_30m | 30mg or more Iron consumption on study completion |
| Sodium (Na)-daily diet consumption | Na_5_4000m | 4000mg or more consumption on study completion |
| K daily diet consumption -completion | K_5_5000m | 5000mg or more consumption on study completion |
| K-daily diet consumption completion | K_5_4000 | 4000mg or more consumption on study completion |
| K daily diet consumption -completion | K_5_3000m | 3000mg or more consumption RDA on study completion |
| Methotrexate (MTX) dose mg per week | MTX_1_16m | 16mg or more weekly dose baseline for more disease activity |
| MTX use | MTX_1_yes | use of MTX at baseline |
| MTX+SZP (sulfasalazine) consumption | MTX_SZP_1_Yes | use of MTX SZP at baseline |
| Prednisolone (Pred) daily dose mg | Pred_1_6m | 6mg or more daily dose of prednisolone at baseline, more active dis |
| Pred use | Pred_1_Yes | Prednisolone use at baseline |
| Non-steroidal anti- inflammatory use (NSAID) | NSAID_1_Yes | NSAID use at baseline |
| Combo use | Combo_1_Yes | Combination use of DMARD at baseline |
| Combo +Pred use | Combo_P_1_Yes | Combination DMARD plus prednisolone at baseline |
| MTX use on completion | MTX_5_Yes | MTX use on study completion |
| MTX dose use on completion | MTX_5_16m | MTX dose 16mg or more on study completion |
| Pred use on completion | Pred_5_Yes | Pred use on study completion |
| Pred dose on completion | Pred_5_6m | Pred dose 6mg or more daily on study completion |
| Combo Use | Combo_5-Yes | Combination DMARD on completion |
| NSAID use on completion | NSAID_5_Yes | NSAID use on completion |
| **Pain on completion** | **Pain_5_4l** | **Dependent Variable : pain less than 4 cm on VAS on study completion** |
| **Disease activity score using ESR less than 3.2** | **DAS28_5_low** | **Dependent Variable: DAS28 low disease or remission on study completion** |
| **Note: ; n: number; m:more;l:less; K: potassium; Combo: combination; MTX: methotrexate; Pred: prednisolone; SZP: sulfasalazine;** | | |

Table 7: Odds Ratio (Association) of Diet Variable and Nutrients with Dependent Variable using Univariate analysis (Z test): A Randomized Assessor Blind three Arm Controlled Diet Intervention Study of Symptomatic Rheumatoid Arthritis (RA) (n=172 patients) of 16 Weeks Duration.

| **Dependent Variable/ Diet related variable** | **Pain change MCID on study completion** | | **Pain VAS on study completion less than 4 cm (VAS)** | | **DAS28 score on study completion less than 3.2** | |
| --- | --- | --- | --- | --- | --- | --- |
|  | **OR** | **‘Z’value^€^** | **OR** | **‘Z’ value^€^** | **OR** | **‘Z’ value^€^** |
| K-arm | 2.0134* | 2.0069 | 1.5450 | 1.2749 | 1.2613 | 0.6474 |
| K-diet-arm | 0.9300 | -0.2061 | 0.9285 | -0.2152 | 1.1229 | 0.3203 |
| SRK-1_3.5l | 0.8467 | -0.4363 | 0.6705 | -1.0712 | 1.3344 | 0.7539 |
| URK-1_40m | 0.5481 | -1.8163 | 0.4711* | -2.3234 | 1.3900 | 0.9675 |
| Energy-1_2700m | 0.4573* | -2.2719 | 0.6638 | -1.2159 | 0.5944 | -1.4589 |
| Prot-1_80m | 0.4805 | -1.9691 | 0.7218 | -0.8948 | 0.4993 | -1.8144 |
| Zn-1_15m | 1.1794 | 0.4254 | 1.0340 | 0.0882 | 0.4639 | -1.9244 |
| VitC-1_130m | 0.7054 | -0.9959 | 0.5611 | -1.6852 | 1.3828 | 0.8996 |
| Iron-1_30m | 1.5119 | 1.2419 | 1.2012 | 0.5628 | 0.6375 | -1.3151 |
| K-1_2200l | 1.2631 | 0.6756 | 1.2777 | 0.7245 | 0.8308 | -0.5212 |
| Energy-5_2700m | 1.3806 | 0.8907 | 0.9775 | -0.0640 | 0.9246 | -0.2103 |
| Prot-5_80m | 0.7950 | -0.6114 | 1.0017 | 0.0047 | 0.5430 | -1.5830 |
| Zn-5_15m | 1.8395 | 1.7396 | 2.0192* | 2.0498 | 0.6301 | -1.2815 |
| Calcium__800m | 1.6071 | 1.3102 | 1.5000 | 1.1443 | 0.7070 | -0.9308 |
| VitC-5_130m | 0.8588 | -0.3807 | 0.7428 | -0.7599 | 1.0715 | 0.1681 |
| Iron-5_30m | 1.6898 | 1.5761 | 1.6532 | 1.5436 | 0.7166 | -0.9732 |
| Na-5_4000m | 3.3526* | 3.4002 | 2.5961* | 2.7405 | 0.9427 | -0.1611 |
| K-5_5000m | 3.8911* | 3.5939 | 2.5909* | 2.5737 | 0.9382 | -0.1638 |
| K-5_4000m PP | 1.8417 | 1.7864 | 1.6561 | 1.5083 | 0.9448 | -0.1614 |
| K-5_3000m | 1.8864 | 1.8889 | 1.5277 | 1.2891 | 0.9388 | -0.1824 |
| Note: n=number; OR: Odds Ratio and testing with population OR=1; €: Estimated after ‘log’ transformation;; *: Statistically Significant as ‘Z’ value is either greater than 1.96 or smaller than -1.96 and therefore included in ‘Logistic Model’; several variables dummy (binary) coded as per investigator discretion and shown in Table 3; m:more; l:less; MCID: minimum clinically important difference (for pain VAS = 1 cm) | | | | | | |

Table 8: Logistic regression models (with stepwise forward) in a randomized controlled diet intervention study of symptomatic rheumatoid arthritis (RA) to determine predictors of low pain (4 cm or less on VAS) at study completion (16 weeks): Shows variables (predictors) with significant regression coefficients (Odds ratio)as output in 4 Models

| **Dependent**  **Variable** | **Group Independent Variables and Method** | **R2** | **Predictor (Odds Ratio)** |
| --- | --- | --- | --- |
| Pain VAS less than 40 cms on study completion | METHOD=ENTER:age_40less,RA>5years, tobacco, menopause, BMI_ 25m,JCPT1_7m,JCSW1_2m,HAQ1_6m,PGA1_3m,P.A.D1_3m, GHA1_60less, EMS1_30m,RAPS1_60m,DAS1_5.1m, SF36P1_40l,SF36M1_40l,ESR1_50m,CRP1_12m,RF1_120m,MTX1_16m,MTX1_yes,MTX_SZP1,yes, PRED1_Yes, Pred1_6m,NSAID1_Yes, Combo1_Yes, ComboP1_Yes,MTX5_16more,MTX5Yes, RED5_Yes,Pred5_6m, NSAID5_Yes, Combo5_yes,SrCort1, K_Arm ,K5_diet arm,SRK1_40ml,UrineK1_40m, ENERGY1_2700m,Prot1_80m, , ZINC1_15m,VITC1_130m, IRON1_30m, K1_2200l,ENERGY5_2700M,Prot5_80m,ZINC5_15m,CALCIUM5_800M, VITC5_130m, IRON5_30m,Na5_4000 m, K5_5000m,K5_4000m, K5_3000m | 60.7 | Menopause (0.138), HAQ1_6m  (0.225), GHA1_60less (0.129),  DAS28_1_5more (12.51), RF1_120 more (4.65), MTX1_yes (108.09),  Pred5_6more(0.055), K5diet arm  (9.58), K5_5000m (20.893) |
|  | METHOD=STEPWISE FORWARD; ALL VARIABLES AS ABOVE IN THE EQUATION; 6 steps to achieve optimum outcome | 32.9 | RA duration >5years (0.29), HAQ1_6more(0.346**), MTX1_yes**  **(16.096)**, MTX_SZP1_yes (0.078)  Pred5_6m (0.327), **K5_5000m**  **(2.876)** |
|  | METHOD=ENTER (Selected variables): age_40less,RA>5 years, menopause JCSW_1_2m P.A.D_1_3m, Pred_5_6m, K_Arm,ENERGY_1_2700more , Na_5_4000more, K_5_5000more | 33 | MTX1_yes (2.818), UrineK1_40 more, (0.42), RA duration > 5 year  (0.341), HAQ1_6more (0.503),  Menopause (0.51), GHA1_60less (0.376), ZINC5_15more (1.941) |
|  | METHOD= STEPWISE FORWARD; ALL THE ABOVE SELECTED VARIABLES IN THE EQUATION; 4 steps. | 25.3 | RA duration>5years (0.295), HAQ1 _6more(0.376), MTX1_yes (2.498),  **K5_5000more (3.145)** |
| Note: All models achieved good fit; n: number; R2:percent of the variation explained by the predictors ,as per the method of Nagelkerke; See Table 6 for abbreviations | | | |
